# Change in the trend of long-term care service usage following COVID-19 pandemic in Japan: a survey using nationwide statistical summary in 2018-2021

**DOI:** 10.1101/2021.12.15.21267784

**Authors:** Kenichiro Sato, Yoshiki Niimi, Takeshi Iwatsubo, Shinya Ishii

## Abstract

**Aim:** Social restriction due to coronavirus disease 2019 (COVID-19) pandemic forced long-term care (LTC) service users to refrain from using services as before, of which degree of change we aim to evaluate in this study.

**Methods:** We retrospectively analyzed publicly-distributed nationwide statistics summarizing the monthly number of public LTC insurance users in Japan in the period between April 2018 and March 2021. The degree of decline was quantified as odds ratio (OR), where the ratio of a certain month to the reference month was divided by the ratio in the previous year.

**Results:** The use of LTC services showed unimodal serial change: it started to decline in March 2020 and reached its largest decline in May 2020, which had insufficiently recovered even as of late 2020. The degree of decline was specifically large in services provided in facilities for community-dwelling elderly individuals (adjusted OR 0.719 (95%CI: 0.664 ∼ 0.777) in short-stay services and adjusted OR 0.876 (95%CI: 0.820 ∼ 0.935) in outpatient services) but was non-significant in other types of service, including those provided for elderly individuals living in nursing homes.

**Conclusions:** Current study showed that community-dwelling elderly individuals who had used outpatient or short-stay services were the segments which were specifically affected by the COVID-19 pandemic in 2020 Japan. It underlines the need for further investigation for the medium- or long-term influence on the mental and physical health of these LTC service users as well as their family caregivers.

## Introduction

As a result of the global outbreak of the new coronavirus (SARS-CoV-2) infection (COVID-19), the COVID-19 pandemic had affected the lives of citizens and their social and economic activities to a great extent [Karako 2021]. Aiming to reduce the risk of infection, we have been forced to adopt changing our lifestyles and behaviors [Arai 2021], such as social distancing [Cato 2020], wearing masks, refraining from unnecessary going-out, working from home, and so on.

From early on the COVID-19 epidemic, it was known that older individuals have a higher risk of severe illness or mortality by COVID-19 infection [GGI guideline 2020], which might have made elderly individuals more likely to accept social restrictions than younger individuals did [Muto 2020]. On the other hand, prolonged social restrictions had been in turn concerned for their negative impact on the mental and physical health of the elderly individuals [Armitage 2020; Plagg 2020].Previous studies have reported several kinds of unfavorable consequences of the pandemic [Loyola 2020], such as a decline in physical activity [Tison 2020; Suzuki 2020; Yamada 2020; Ono 2021] due to refraining from going out, increased social isolation [Sugaya 2021] (e.g., decreased opportunities to connect face-to-face with relatives and friends, restricted family visits at long-term care facilities, or isolation of residents in their private rooms [Chu 2020; Wang 2020; Bethel 2021]), increased anxiety and depression during social isolation [Loyola 2020], loneliness [Salman 2021], or the decreased well-being [Suzuki 2020]. The loneliness, social isolation, and decreased social participation are in general associated with the risk of dementia and cognitive deterioration [Ross 2018; Lara 2019; Sundstrom 2019], and indeed serial cognitive deterioration due to social isolation [Noguchi 2021] or a decline in cognitive fitness during the period when the nationwide state of emergency (SoE) had been declared [Makizako 2021] were reported so far.

The provision system of long-term care (LTC) services, which play a critical role in maintaining the daily lives and functions of elderly individuals [Maruta 2019], was greatly affected by the COVID-19 pandemic, too. In Japan, LTC services are to be provided to individual users according to the needs of daily life by taking into account their physical and cognitive functions [Maruta 2019], so that the gap in utilization of LTC services between before and during pandemic represents at least one part of the above-mentioned decrease in physical activity and social participation by itself. In other words, examining a detailed aspect of the degree and characteristics of change in the use of LTC services will be one of the alternative measures for understanding the impact of the COVID-19 pandemic on the mental and physical health of elderly people. In addition, since the care burden of a declined use would be partly passed on the caregiving burden of their family members [Sugawara 2021], examining the change in LTC service use by elderly individuals following pandemic will also be helpful in understanding the actual change in caregiving burdens [Mazzi 2020; Altieri 2021; Borelli 2020] for their family members.

In this study, we aim to evaluate the degree of decline in the use of LTC services by older individuals in Japan following the pandemic, using nationwide statistics which is opened to the public every year. Although the database only includes the total monthly number of each service user, it has an advantage in terms of its high accessibility and analyzability to grasp an overview of changes in the LTC service use following pandemic and to provide a basis for further research.

## Methods

### Data acquisition

We retrospectively analyzed nationwide statistics summarizing the monthly number of public LTC insurance users in Japan in the period between April 2018 and March 2021, which has been opened to the public by the Ministry of Health, Labour and Welfare (MHLW), Japan. Annual summary statistics has been opened to the public every year on its website (https://www.mhlw.go.jp/toukei/list/45-1b.html), and it includes the monthly count of all users across Japan by each service without distinguishing by the prefecture of residence.

### About Japanese LTC insurance system

The public LTC insurance system in Japan provides various types of services to community-dwelling people and to residents in nursing homes. In general, every community-dwelling people 65 years or older, or those aged 40-64 with a disability due to some specific diseases, can use several types of LTC services simultaneously with an arbitrary combination as per their certified need of support/care. The upper limit in the monthly amount of available services is regulated based on the ‘degree of care required’, which is mainly determined based on the two-axes disability of individual users - physical disability and cognitive decline. The classification in physical disability and cognitive decline is to be made especially in terms of dependence on care, which is determined by users’ home doctors when applying certification (either new or updated application). And the ‘degree of care required’ is determined by the municipal judging committee based on the pre-determined physical disability and cognitive decline. Certification needs to be updated regularly (e.g., every 6-36 months), although every certified user does not always have to use any services. In 2019, more than 6 million people in Japan had been certified of any degree. Determined ‘degree of care required’ may slightly differ by each municipality, reflecting regional circumstances even with the same level of disabilities, but the amount of services available in each ‘degree of care required’ level is common throughout the country.

### LTC services

Various LTC services for community-dwelling people and individuals living at nursing facilities are categorized into a few service types depending on their features including the place of service provided, as follows (Table 1): ‘home-visit’ services where service providing staff visits users’ own house and the services including care, nursing, or rehabilitation are provided there. ‘Outpatient’ services where each user visits service provider facilities and receives LTC services including care (so-called ‘day-service’) or rehabilitation (so-called ‘day-care’). In ‘rental’ service, LTC users can rental specific equipments for welfare, such as walking sticks, walkers, or wheelchairs. ‘Short-stay’ services offer users admit facilities for a short-term recuperation. ‘Guidance’ service provides users some specialised management or guidance (but not including medical treatment), which is made by home-visiting doctors, dentists, pharmacists, nurses, or dietitians. ‘Designated facility’ service provides daily life care for individuals living at designated facilities. ‘Facility’ is another type of service provided for those living in care facilities or nursing homes. And ‘community-based’ services where the combination of various kinds of services are provided as a one-stop service.

**Table 1.** Name of services analyzed and their types

| Type | Code | Name of service | Preventive |
| --- | --- | --- | --- |
| Home-visit | 11 | Home-visit care |  |
|  | 12 | Home-visit bathing |  |
|  | 13 | Home-visit nursing |  |
|  | 14 | Home-visit rehabilitation |  |
|  | 63 | Home-visit nursing [preventive] | ✓ |
|  | 64 | Home-visit rehabilitation [preventive] | ✓ |
| Outpatient | 15 | Outpatient care (day-service) |  |
|  | 16 | Outpatient rehabilitation (day-care) |  |
|  | 66 | Outpatient rehabilitation [preventive] | ✓ |
| Rental | 17 | Rental of welfare equipment |  |
|  | 67 | Rental of welfare equipment [preventive] | ✓ |
| Short-stay | 21 | Short-stay for daily care |  |
|  | 22 | Short-stay for recuperation (at care facility) |  |
| Guidance | 31 | Management guidance for in-home care |  |
|  | 34 | Management guidance for in-home care [preventive] | ✓ |
| Designated facility | 33 | Daily life care at designated facilities |  |
|  | 35 | Daily life care at designated facilities [preventive] | ✓ |
| Community-based | 76 | Periodic visits and occasional home-visit care/nursing |  |
|  | 78 | Community-based outpatient care |  |
|  | 72 | Outpatient care for dementia patients |  |
|  | 72 | Multifunctional in-home care |  |
|  | 32 | Group home care for dementia patients |  |
|  | 54 | Community-based special nursing homes |  |
|  | 75 | Multifunctional in-home care [preventive] | ✓ |
| Facility | 51 | Welfare facility (special nursing homes) |  |
|  | 52 | Health care facility |  |

The English name of services listed above are not always official ones since they are simplified by the authors for clarity.

### Quantifying change in service usage

All the following data processing and statistical analyses were conducted by using R version 3.6.3 and its packages. Since use in service may be influenced by seasonal fluctuation as well as a longer trend in the recent few years, we quantify the degree of use in month *M* by taking odds ratio (OR) and its 95% confidence interval (CI), which are defined by the following equations (a,b,c,∈ ℕ):

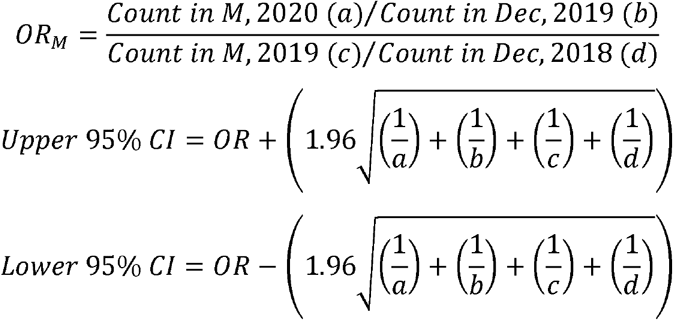

*OR*_*M*_ means the degree of change in the user count in month *M* of 2020 compared to the same month in the previous year (*M* = January ∼ November) and the reference month (= December) in the previous year. Lower 95% CI > 1 shows the increased usage, and upper 95% CI < 1 means the decreased usage of the service. OR of service *s* across the country is specified as *OR*_*s,M*_. In total, 26 service subgroup ORs were obtained in each month (Jan ∼ Nov 2020).

### Meta-regression

For all *OR*_*s*_ in a fixed month *M*, we evaluate whether factors related to the type of service may bring heterogeneity as evaluated by I^2^ value and p-value in Q-test. We conduct multiple meta-regression analysis, of which equation is described as follows:

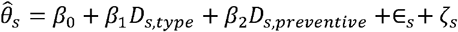

Effect size 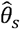 of subgroup *s*, which corresponds to the name of service, is regressed by the subgroups’ covariates: *D*_*s,type*_ is the dummy variable as to the type of service the service *s* belongs to (Table 1, left-most column), and *D*_*s, preventive*_ is the dummy variable whether the service *s* is preventive service or not (Table 1, right-most column). And ε_*s*_ is the sampling error, and ζ_*s*_ is the random effect. Obtained β_1_ and β_2_ for each month *M* is summarized to overview the serial changes of each feature.

## Results

### Serial trend: overview

The serial trend in the number of monthly users in each of 26 services is plotted in Figure 1. Figure 1B shows the normalized series in Figure 1A, which was obtained by dividing by the mean number of users in 2019. Visually, many of the series started to decline mildly 1-2 months before April-May 2020, which corresponds to vertical red bands in the figure, then significantly decreased in April and May 2020, during which the first nationwide SoE had been declared. After June 2020, the series gradually recovered, but the degree of recovery has not been sufficient in some service series even as of March 2021.

**Figure 1.**
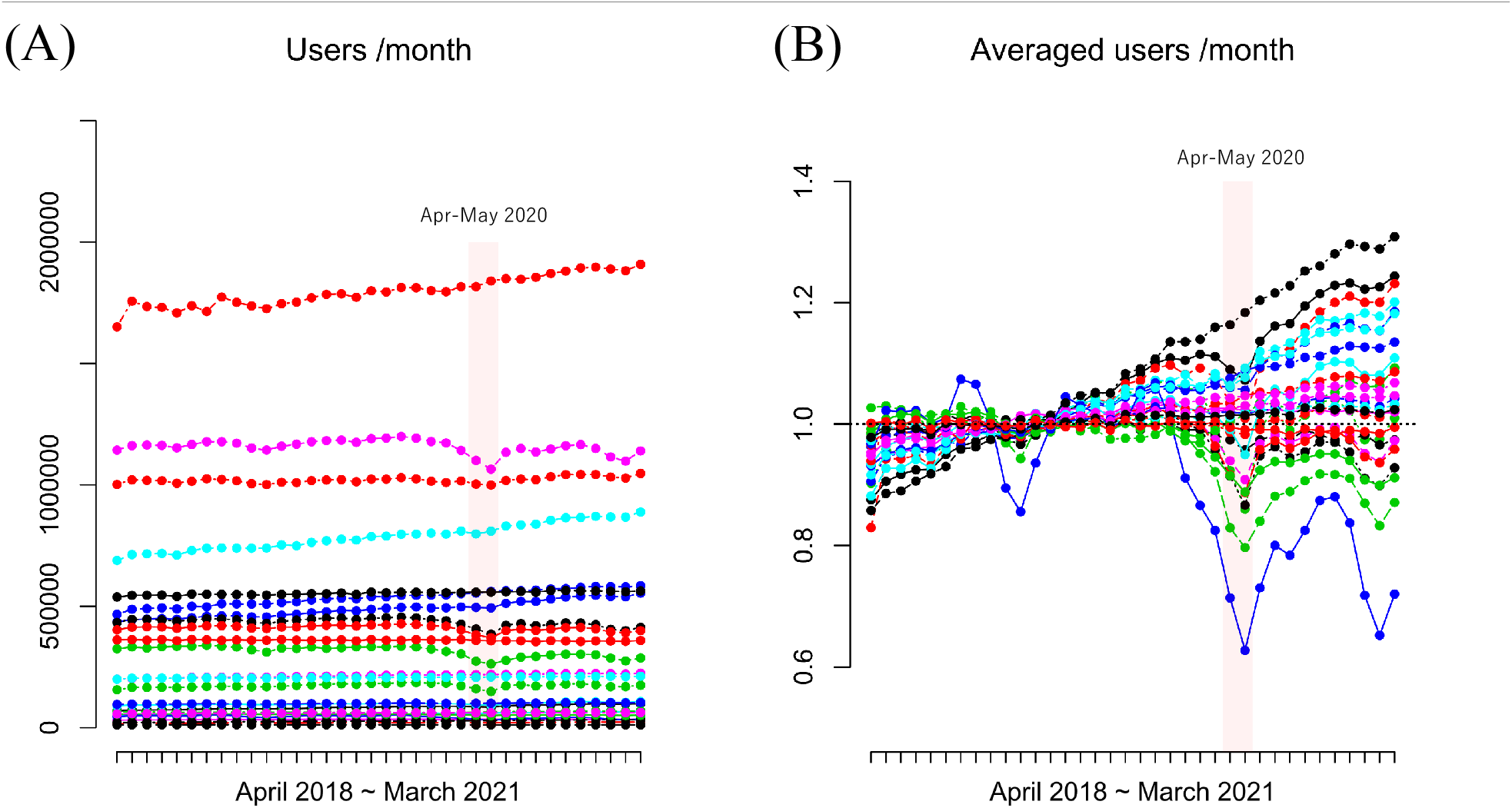
The serial trend in the number of monthly users in each of 26 services

### Serial change by the type of service

We then obtained OR at each month for individual services, as visualized in Figure 2, where each plot shows serial ORs from January to November 2020. Figure 2A shows the serial change of ‘home-visit’ services, and most of them except for ‘home-visit bathing’ showed a mild decline in May 2020. Figure 2B shows the serial change of ‘outpatient’ services, and they showed a moderate decline in May 2020. Figure 2C, 2E, 2F, and 2H show serial changes of ‘rental’, ‘guidance’, ‘designated’, and ‘facility’ services, respectively, and they showed no significant or only slight changes in May 2020. Figure 2D shows the serial change of ‘short-stay’ services, and they showed the largest decline in May 2020. Figure 2G shows the serial change of ‘community-based’ services, where many of them showed no significant changes. In addition, outpatient services and short-stay services had insufficient recovery even in late 2020.

**Figure 2.**
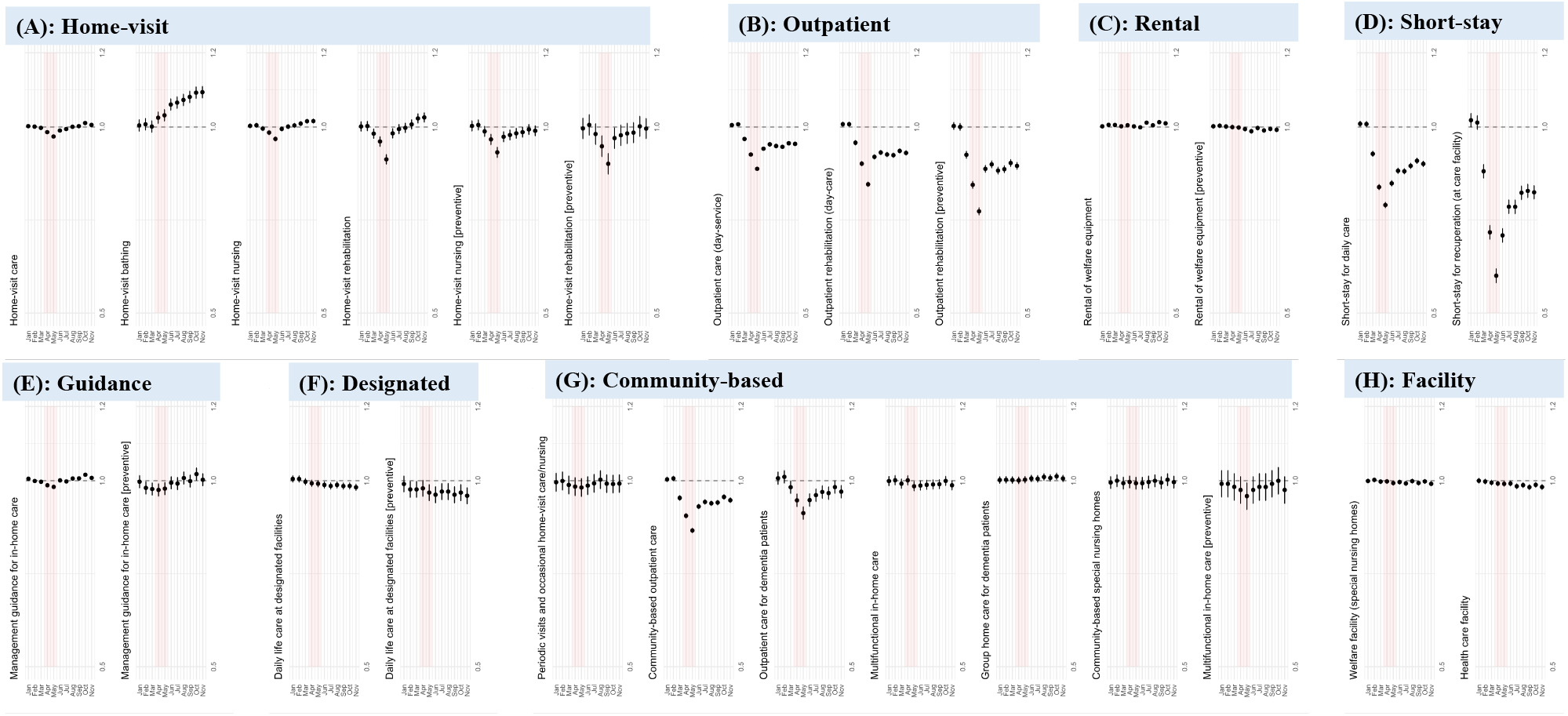
OR in series Serial change from January to November 2020. Red bands denote April-May, when the nationwide state of emergency had been declared.

### Meta-regression

We now conducted meta-regression analysis: Figure 3 means the serial trend in the obtained coefficients from January to November 2020. Changes in the home-visit services were non-significant as partly represented by the series of the intercept term (a). Compared to the series of home-visit services, outpatient services (b) showed a moderate level of change in April-May, while short-stay services (d) showed ther severest change in April-May. No other types of service showed significant decline in April-May. In addition, no significant change was also observed in the case of prevention services (i).

**Figure 3.**
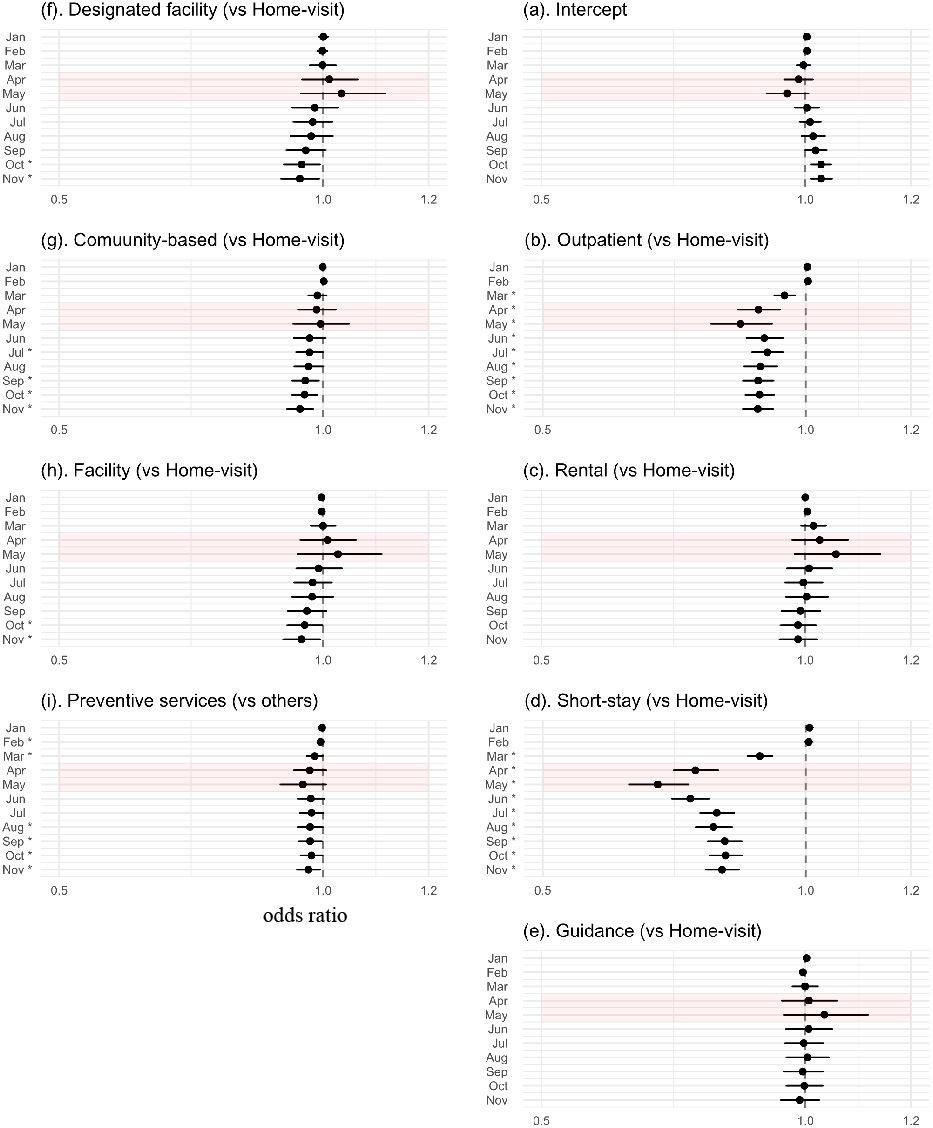
Serial change in the obtained coefficients by meta-regression Asterisk (*) means a significant decrease in the month.

### Ethics

This study has been approved by the University of Tokyo Graduate School of Medicine institutional ethics committee (ID: 11628-(3)). Ethical approval is not required for this type of study. This work was conducted in accordance with the ethical standards, laid down in the 1964 Helsinki declaration.

## Discussion

In this study, we quantitatively evaluated the degree of decline in the use of LTC services across Japan. We identified that the use of LTC services showed unimodal serial change, especially in outpatient and short-stay services for community-dwelling individuals: it started to decline in March 2020 and reached its largest decline in May 2020, which had insufficiently recovered even as of November 2020. There was no significant change in the use of home-visiting services or services for residents in nursing homes. The current study showed that community-dwelling elderly individuals who had previously used outpatient or short-stay services were the segment who had been specifically affected by the COVID-19 pandemic. Despite the shortness that this study used the database which only equipped with the total monthly number of each service user, it still has an advantage in terms of its high accessibility and analyzability to grasp an overview of changes in the LTC service use following the pandemic, thereby providing a basis for further research: medium- or long-term influence on the mental health of these LTC service users as well as their family caregivers.

There were some additional considerations that can be suspected from the serial change in users. The degree of decline was not steep in response to the declaration of SoE since usage decline had already started as of March. This means SoE was not a primary factor that induced withdrawal of usage, but rather an additional factor in deteriorating the decline in usage, supporting our experiences. In addition, the recovery in short-stay and outpatient service users remained insufficient as of November, possibly reflecting the continuous set of a limit in the number of available users at once to reduce the risk of infection. Meanwhile, it is uncertain where the decline of use in late 2020 had been passed on: the declined amount of service was shifted to the use of other types of LTC services or to the caregiving burden of their family members?

We can enumerate various kinds of scenarios in which individual community-dwelling LTC users did not receive LTC services as before the pandemic, such as follows: LTC facilities providing services were temporarily closed due to nosocomial outbreak (or ‘cluster’) [Iritani 2020], LTC users were required to self-isolate in their homes because they were considered as a close contact since their family member living together became COVID-19 positive, LTC users’ excessive concerns or anxieties of infection prevented them from using LTC services, or some LTC users had a lower need to continue LTC services because they had relatively been independent since previously [Hirose 2021]. What we need to note is that the reason for the decline in use is not only due to factors on the demand side of the service, but also includes factors on the supply side of the service such as temporary closure due to the spread of infection among facility staff and users. It would be difficult to distinguish between these factors in a data-basis approach, even when we had used a comprehensive nationwide claims database.

In response to the COVID-19 pandemic to lower the risk of infection among LTC users and to make users/providers ease to use/provide LTC services, there were several kinds of exceptional measures made in the operation policy of the LTC insurance system. For example, in order to prevent crowding in service facilities in the purpose of reducing the risk of infection, home-visit services were temporarily permitted to substitute outpatient services (office correspondence from MHLW: https://www.mhlw.go.jp/content/000605436.pdf). And an earlier study did report reduced usage in outpatient services but maintained usage in home-visit services [Ito 2021]. The current study showed that the above attempt to maintain the provisioning system of home-visit services for community-dwelling elderlies had been sustained to a considerable degree. No significant change in the users of home-visiting services means two possible cases: some proportion of declined users for outpatient or short-stay services were transferred by using home-visiting service which declined mildly, or the use of outpatient or short-stay services simply declined but there was no decline in the use of home-visit service. The monthly proportion of new users will be a key to distinguishing these possible patterns, which needs to be evaluated in future research. Currently, we suspect that it does not seem to clearly indicate the tendency where people who had been using outpatient or short-stay services have switched to the home-visit services as a temporary alternatives. This point is consistent with the previous literature [Sugawara 2021], which discussed that the replacement of outpatient services with home-visit services was not conducted insufficiently.

This study includes some limitations. First, the current study is based on the summary statistics across the country, without distinguishing the difference in each prefecture in terms of the baseline service usage or the degree of disruption by the COVID-19 pandemic. So that the degree of decline obtained in this study is more likely to reflect the changes in metropolitan areas with a larger populations such as Tokyo or Osaka, and the changes in prefectures with relatively smaller populations are at risk of being masked. Second, we referred to the monthly count of users in the period from December 2018 to December 2020 in order to quantify the degree of change, this means the serial trend in a much longer level (e.g., 3-5 years or so) cannot be incorporated, leading to a risk of inaccurate estimation in change. Using interrupted time-series analysis (ITSA) may help to this point, although the ITSA may also have some disadvantages in that the emergence of decline was not sudden. And third, it is unclear from this data alone whether the change in users is truly related to the COVID-19 pandemic or whether it is just a pseudo-correlation that was originally caused by another event.

Using a nationwide LTC claims database might be of help to examine further detailed changes following the pandemic. Specifically, incorporating difference by municipality is important. In addition to the difference in the baseline populations by prefecture, the difference in the amount of COVID-19 patients per population by prefecture should also be considered. For example, on May 25 of 2020 when the first nationwide SoE was canceled, there were 5,160 cumulative cases in Tokyo prefecture (= 3.7 per 100,000 population) and 0 cumulative cases in Iwate prefecture (= 0.0 per 100,000 population) (https://covid19.mhlw.go.jp). The government of Japan had specified certain 12 prefectures which have a large amount of COVID-19 cases or whose COVID-19 cases increased with fast speed as ‘designated prefectures under specific cautions’, where social restrictions were slightly harder than in other prefectures. These specific prefectures included Hokkaido, Ibaraki, Saitama, Chiba, Tokyo, Kanagawa, Ishikawa, Gifu, Aichi, Osaka, Hyogo, and Fukuoka prefectures. Although the provision of LTC services had not been directly restricted in these prefectures, its provision and usage may have been affected to any extent indirectly, so that we would also need to incorporate the factor of ‘designated prefectures’ into the analysis.

## Data Availability

The database is opened to public and can be used by anyone who access to the MHLW's website (https://www.mhlw.go.jp/toukei/list/45-1b.html).

## Acknowldgement

This study was supported by JSPS KAKENHI Grant number JP21K20891.

## Data availability

The database is opened to public and can be used by anyone who access to the MHLW’s website (https://www.mhlw.go.jp/toukei/list/45-1b.html). The authors obtained the data on December 3, 2021.

## References

1. Karako K, Song P, Chen Y, Tang W, Kokudo N. Overview of the characteristics of and responses to the three waves of COVID-19 in Japan during 2020-2021. Biosci Trends. 2021 Mar 15;15(1):1–8. doi: 10.5582/bst.2021.01019. Epub 2021 Jan 29. PMID: 33518668.

2. Cato S, Iida T, Ishida K, Ito A, McElwain KM, Shoji M. Social distancing as a public good under the COVID-19 pandemic. Public Health. 2020 Nov;188:51–53. doi: 10.1016/j.puhe.2020.08.005. Epub 2020 Aug 13. PMID: 33120232; PMCID: PMC7425541.

3. Arai Y, Oguma Y, Abe Y, Takayama M, Hara A, Urushihara H, Takebayashi T. Behavioral changes and hygiene practices of older adults in Japan during the first wave of COVID-19 emergency. BMC Geriatr. 2021 Feb 24;21(1):137. doi: 10.1186/s12877-021-02085-1. PMID: 33627073; PMCID: PMC7903371.

4. Japan Geriatrics Society Subcommittee on End-of-Life Issues and New Coronavirus Countermeasure Team, Kuzuya M, Aita K, Katayama Y, Katsuya T, Nishikawa M, Hirahara S, Miura H, Yanagawa M, Arai H, Iijima K, Okochi J, Kozaki K, Yamaguchi Y, Rakugi H, Akishita M. The Japan Geriatrics Society consensus statement “recommendations for older persons to receive the best medical and long-term care during the COVID-19 outbreak-considering the timing of advance care planning implementation”. Geriatr Gerontol Int. 2020 Dec;20(12):1112–1119. doi: 10.1111/ggi.14075. Epub 2020 Nov 2. PMID: 33137849.

5. Muto K, Yamamoto I, Nagasu M, Tanaka M, Wada K. Japanese citizens’ behavioral changes and preparedness against COVID-19: An online survey during the early phase of the pandemic. PLoS One. 2020 Jun 11;15(6):e0234292. doi: 10.1371/journal.pone.0234292. PMID: 32525881; PMCID: PMC7289361.

6. Armitage R, Nellums LB. COVID-19 and the consequences of isolating the elderly. Lancet Public Health. 2020 May;5(5):e256. doi: 10.1016/S2468-2667(20)30061-X. Epub 2020 Mar 20. PMID: 32199471; PMCID: PMC7104160.

7. Plagg B, Engl A, Piccoliori G, Eisendle K. Prolonged social isolation of the elderly during COVID-19: Between benefit and damage. Arch Gerontol Geriatr. 2020 Jul-Aug;89:104086. doi: 10.1016/j.archger.2020.104086. Epub 2020 May 3. PMID: 32388336; PMCID: PMC7196375.

8. Sepúlveda-Loyola W, Rodríguez-Sánchez I, Pérez-Rodríguez P, Ganz F, Torralba R, Oliveira DV, Rodríguez-Mañas L. Impact of Social Isolation Due to COVID-19 on Health in Older People: Mental and Physical Effects and Recommendations. J Nutr Health Aging. 2020;24(9):938–947. doi: 10.1007/s12603-020-1469-2. PMID: 33155618; PMCID: PMC7597423.

9. Tison GH, Avram R, Kuhar P, Abreau S, Marcus GM, Pletcher MJ, Olgin JE. Worldwide Effect of COVID-19 on Physical Activity: A Descriptive Study. Ann Intern Med. 2020 Nov 3;173(9):767–770. doi: 10.7326/M20-2665. Epub 2020 Jun 29. PMID: 32598162; PMCID: PMC7384265.

10. Suzuki Y, Maeda N, Hirado D, Shirakawa T, Urabe Y. Physical Activity Changes and Its Risk Factors among Community-Dwelling Japanese Older Adults during the COVID-19 Epidemic: Associations with Subjective Well-Being and Health-Related Quality of Life. Int J Environ Res Public Health. 2020 Sep 10;17(18):6591. doi: 10.3390/ijerph17186591. PMID: 32927829; PMCID: PMC7557874.

11. Yamada M, Kimura Y, Ishiyama D, Otobe Y, Suzuki M, Koyama S, Kikuchi T, Kusumi H, Arai H. Effect of the COVID-19 Epidemic on Physical Activity in Community-Dwelling Older Adults in Japan: A Cross-Sectional Online Survey. J Nutr Health Aging. 2020;24(9):948–950. doi: 10.1007/s12603-020-1424-2. PMID: 33155619; PMCID: PMC7597428.

12. Ono T, Kashima M, Asakawa Y. Self-rated Changes of Health Status during Stay-at-home Orders among Older Adults Using the Long-term Care Insurance System of Japan: A Cross-sectional Study. Phys Ther Res. 2021 Apr 1;24(2):170–175. doi: 10.1298/ptr.E10089. PMID: 34532213; PMCID: PMC8419481.

13. Chu CH, Wang J, Fukui C, Staudacher S, A Wachholz P, Wu B. The Impact of COVID-19 on Social Isolation in Long-term Care Homes: Perspectives of Policies and Strategies from Six Countries. J Aging Soc Policy. 2021 Jul-Oct;33(4-5):459-473. doi: 10.1080/08959420.2021.1924346. Epub 2021 May 9. PMID: 33969815.

14. Wang H, Li T, Barbarino P, Gauthier S, Brodaty H, Molinuevo JL, Xie H, Sun Y, Yu E, Tang Y, Weidner W, Yu X. Dementia care during COVID-19. Lancet. 2020 Apr 11;395(10231):1190–1191. doi: 10.1016/S0140-6736(20)30755-8. Epub 2020 Mar 30. PMID: 32240625; PMCID: PMC7146671.

15. Bethell J, Aelick K, Babineau J, Bretzlaff M, Edwards C, Gibson JL, Hewitt Colborne D, Iaboni A, Lender D, Schon D, McGilton KS. Social Connection in Long-Term Care Homes: A Scoping Review of Published Research on the Mental Health Impacts and Potential Strategies During COVID-19. J Am Med Dir Assoc. 2021 Feb;22(2):228-237.e25. doi: 10.1016/j.jamda.2020.11.025. Epub 2020 Nov 26. PMID: 33347846.

16. Sugaya N, Yamamoto T, Suzuki N, Uchiumi C. Social isolation and its psychosocial factors in mild lockdown for the COVID-19 pandemic: a cross-sectional survey of the Japanese population. BMJ Open. 2021 Jul 14;11(7):e048380. doi: 10.1136/bmjopen-2020-048380. PMID: 34261687; PMCID: PMC8282418.

17. Salman D, Beaney T E, Robb C, de Jager Loots CA, Giannakopoulou P, Udeh-Momoh CT, Ahmadi-Abhari S, Majeed A, Middleton LT, McGregor AH. Impact of social restrictions during the COVID-19 pandemic on the physical activity levels of adults aged 50-92 years: a baseline survey of the CHARIOT COVID-19 Rapid Response prospective cohort study. BMJ Open. 2021 Aug 25;11(8):e050680. doi: 10.1136/bmjopen-2021-050680. PMID: 34433606; PMCID: PMC8390149.

18. Penninkilampi R, Casey AN, Singh MF, Brodaty H. The Association between Social Engagement, Loneliness, and Risk of Dementia: A Systematic Review and Meta-Analysis. J Alzheimers Dis. 2018;66(4):1619–1633. doi: 10.3233/JAD-180439. PMID: 30452410.

19. Lara E, Caballero FF, Rico-Uribe LA, Olaya B, Haro JM, Ayuso-Mateos JL, Miret M. Are loneliness and social isolation associated with cognitive decline? Int J Geriatr Psychiatry. 2019 Nov;34(11):1613–1622. doi: 10.1002/gps.5174. Epub 2019 Jul 25. PMID: 31304639.

20. Sundström A, Adolfsson AN, Nordin M, Adolfsson R. Loneliness Increases the Risk of All-Cause Dementia and Alzheimer’s Disease. J Gerontol B Psychol Sci Soc Sci. 2020 Apr 16;75(5):919–926. doi: 10.1093/geronb/gbz139. PMID: 31676909; PMCID: PMC7161366.

21. Noguchi T, Kubo Y, Hayashi T, Tomiyama N, Ochi A, Hayashi H. Social Isolation and Self-Reported Cognitive Decline Among Older Adults in Japan: A Longitudinal Study in the COVID-19 Pandemic. J Am Med Dir Assoc. 2021 Jul;22(7):1352-1356.e2. doi: 10.1016/j.jamda.2021.05.015. Epub 2021 May 21. PMID: 34107288.

22. Makizako H, Nakai Y, Shiratsuchi D, Akanuma T, Yokoyama K, Matsuzaki-Kihara Y, Yoshida H. Perceived declining physical and cognitive fitness during the COVID-19 state of emergency among community-dwelling Japanese old-old adults. Geriatr Gerontol Int. 2021 Apr;21(4):364–369. doi: 10.1111/ggi.14140. Epub 2021 Feb 11. PMID: 33576180; PMCID: PMC8013798.

23. Maruta M, Tabira T, Makizako H, Sagari A, Miyata H, Yoshimitsu K, Han G, Yoshiura K, Kawagoe M. Impact of Outpatient Rehabilitation Service in Preventing the Deterioration of the Care-Needs Level Among Japanese Older Adults Availing Long-Term Care Insurance: A Propensity Score Matched Retrospective Study. Int J Environ Res Public Health. 2019 Apr 10;16(7):1292. doi: 10.3390/ijerph16071292. PMID: 30974921; PMCID: PMC6480242.

24. Sugawara S, Nakamura J. Long-term care at home and female work during the COVID-19 pandemic. Health Policy. 2021 Jul;125(7):859–868. doi: 10.1016/j.healthpol.2021.04.013. Epub 2021 Apr 30. PMID: 33994215; PMCID: PMC8084915.

25. Carpinelli Mazzi M, Iavarone A, Musella C, De Luca M, de Vita D, Branciforte S, Coppola A, Scarpa R, Raimondo S, Sorrentino S, Lualdi F, Postiglione A. Time of isolation, education and gender influence the psychological outcome during COVID-19 lockdown in caregivers of patients with dementia. Eur Geriatr Med. 2020 Dec;11(6):1095–1098. doi: 10.1007/s41999-020-00413-z. Epub 2020 Oct 14. PMID: 33052535; PMCID: PMC7556578.

26. Altieri M, Santangelo G. The Psychological Impact of COVID-19 Pandemic and Lockdown on Caregivers of People With Dementia. Am J Geriatr Psychiatry. 2021 Jan;29(1):27–34. doi: 10.1016/j.jagp.2020.10.009. Epub 2020 Oct 22. PMID: 33153872; PMCID: PMC7577876.

27. Borelli WV, Augustin MC, de Oliveira PBF, Reggiani LC, Bandeira-de-Mello RG, Schumacher-Schuh AF, Chaves MLF, Castilhos RM. Neuropsychiatric Symptoms in Patients with Dementia Associated with Increased Psychological Distress in Caregivers During the COVID-19 Pandemic. J Alzheimers Dis. 2021;80(4):1705–1712. doi: 10.3233/JAD-201513. PMID: 33646168.

28. Iritani O, Okuno T, Hama D, Kane A, Kodera K, Morigaki K, Terai T, Maeno N, Morimoto S. Clusters of COVID-19 in long-term care hospitals and facilities in Japan from 16 January to 9 May 2020. Geriatr Gerontol Int. 2020 Jul;20(7):715–719. doi: 10.1111/ggi.13973. PMID: 32634849; PMCID: PMC7361521.

29. Hirose T, Sawaya Y, Shiba T, Ishizaka M, Onoda K, Kubo A, Urano T. Characteristics of patients discontinuing outpatient services under long-term care insurance and its effect on frailty during COVID-19. PeerJ. 2021 Apr 5;9:e11160. doi: 10.7717/peerj.11160. PMID: 33868818; PMCID: PMC8029655.

30. Ito T, Hirata-Mogi S, Watanabe T, Sugiyama T, Jin X, Kobayashi S, Tamiya N. Change of Use in Community Services among Disabled Older Adults during COVID-19 in Japan. Int J Environ Res Public Health. 2021 Jan 28;18(3):1148. doi: 10.3390/ijerph18031148. PMID: 33525441; PMCID: PMC7908432.

